# Insulin genetic clusters and clinical outcomes in Hispanics/Latinos

**DOI:** 10.64898/2026.11.03.26367229

**Authors:** Kaylia M. Reynolds, James P. Lash, Andrew P. Morris, Martha Daviglus, Linda Gallo, Kent D. Taylor, Jiawen Cai, Jinrui Cui, Holly J. Kramer, Yii-Der I. Chen, Daniela Sotres-Alvarez, Jerome I. Rotter, Mark O. Goodarzi, Nora Franceschini

## Abstract

Increased fasting insulin reflects insulin resistance and is as a precursor to type 2 diabetes and a risk factor for kidney, heart and liver diseases and mortality. Cluster-specific fasting insulin polygenic scores (PS) characterize insulin resistance mechanisms based on insulin-related genetic associations with metabolic traits, and they have not been evaluated for liver outcomes. We calculated seven previously developed FI cluster-specific PS, including non-diabetogenic (Preserved Insulin Secretion, Elevated Insulin Secretion, Proinsulin) and diabetogenic clusters (Adiposity-driven Hyperinsulinemia, Visceral Adiposity, Lipodystrophy, Liver/Lipid) in 12,631 participants of the Hispanic Community Healthy Study/Study of Latinos (HCHS/SOL), and tested their association with type 2 diabetes, cardiovascular, kidney and liver outcomes. We also tested the association of FI clusters with indices of insulin homeostasis in additional 1,316 Hispanic individuals. Among HCHS/SOL participants (mean age 46 years, 59% women, 20% with diabetes), we validated associations of FI cluster-specific PS and the 19 traits used to generate clusters. The Lipodystrophy cluster was associated with type 2 diabetes (p-value =3.5 x 10^-5^). In addition, the Adiposity-driven Hyperinsulinemia cluster was associated with the hepatic steatosis index (p-value = 2.2 ×10^−17^) and the fatty liver index (p-value = 1.7 ×10^−14^), while the Liver/Lipid cluster with liver fibrosis (p-value = 1.5 ×10^−4^) and lower kidney function (p-value = 5.2 x 10^-5^). The non-diabetogenic Elevated Insulin Secretion cluster-specific PS was associated with the disposition index (p = 0.0015). These results support insulin resistance mechanisms implicated in type 2 diabetes, liver and kidney outcomes.

---

Diabetes impacts 12% of the United States population, with about 90–95% of cases being type 2 diabetes^1, 2^. In 2024, the age-adjusted prevalence of diagnosed type 2 diabetes in Hispanics/Latinos was 11.8% compared to 7.1% in non-Hispanic white individuals ^1^. Type 2 diabetes is diagnosed at a rate 13% higher in Hispanics/Latinos than the total U.S. adult population ^3^. Hispanics/Latinos are also overrepresented among individuals with a type 2 diabetes diagnosis before 40 years of age ^4^. Type 2 diabetes carries a high burden of mortality, disability, and health care cost in Hispanics/Latinos compared to non-Hispanic white individuals ^5, 6^. Therefore, detection, prevention and treatment of type 2 diabetes is a high priority in this population.

Increases in fasting insulin (FI) have been identified as a precursor to type 2 diabetes development and a risk factor for microalbuminuria, cardiovascular disease, liver disease and mortality ^7–10^. Studies have used genetic data to distinguish type 1 from type 2 diabetes^11^, given different pathophysiology, and to uncover biologic mechanisms underlying type 2 diabetes clinical heterogeneity ^12–14^. A recent study developed genetic clusters for mechanisms related to insulin secretion and sensitivity that precede type 2 diabetes by clustering insulin-related variants based on their association with metabolic traits ^15^. The study identified three clusters related to non-diabetogenic hyperinsulinemia (Preserved Insulin Secretion [Preserved IS], Elevated Insulin Secretion [Elevated IS], Proinsulin clusters) and four diabetogenic clusters (Adiposity-driven Hyperinsulinemia, Visceral Adiposity, Lipodystrophy, Liver/Lipid clusters). Cluster-specific polygenic score (PS) associations with clinical outcomes varied by cluster. The Adiposity-driven Hyperinsulinemia cluster-specific PS and the Lipodystrophy cluster-specific PS were associated with increased hypertension, coronary artery disease (CAD), and chronic kidney disease (CKD), while the Liver/Lipid cluster-specific PS was associated with reduced CAD.

Although the original study tested associations of FI clusters in Hispanics/Latinos with clinical outcomes, these clusters were not validated in Hispanics/Latinos^15^. In addition, several outcomes were not reported including prediabetes states, liver fat and fibrosis, and increased urine albumin. Lastly, PS-specific clusters were not characterized for insulin homeostasis mechanisms. We used a large data of the Hispanic Community Health Study/Study of Latinos (HCHS/SOL) to validate FI cluster-specific PS in Hispanics/Latinos and tested the associations of these clusters with cardiometabolic risk factors, indexes for liver fat and fibrosis, and cardiovascular and kidney outcomes.

Because FI clusters are related to type 2 diabetes mechanisms, we first use a polygenetic score to differentiate participants with type 1 diabetes from type 2 diabetes before testing the association of hyperinsulinemia cluster-specific PS with clinical outcomes. To shed light on the pathophysiology underlying these clusters, we also assessed their association with indices of insulin homeostasis (insulin sensitivity, insulin secretion, insulin clearance, disposition index) in the Mexican American Coronary Artery Disease (MACAD) and Hypertension-Insulin Resistance (HTN-IR) cohorts that have undergone euglycemic hyperinsulinemic clamps and oral glucose tolerance tests.

## Methods

### HCHS/SOL study description

The HCHS/SOL is a multi-center cohort study of 16,415 Hispanics/Latinos, aged 18 to 74 years, recruited from four US field centers (Chicago, Miami, the Bronx, and San Diego) ^16^. The design of the study consisted of a two-stage probability sampling of households in predefined census-block groups This study used the baseline exam (2008 to 2011) and visit 2 exam (2014-2017), which obtained clinical, behavioral, and sociodemographic assessments, and collected fasting blood and spot urine samples. Participants underwent additional survey assessments at follow-up. Serum creatinine was measured using a creatinase enzymatic method traceable to an isotope dilution mass spectrometry reference^17^. Urine albumin (mg/dl) and creatinine (g/dl) were measured using an immunoturbidometric and a creatinase enzymatic method, respectively. HbA1c concentration was measured in EDTA whole blood using a Tosoh G7 Automated HPLC Analyzer. Participants without self-reported diabetes and/or not taking antihyperglycemic medications and/or a fasting plasma glucose ≤ 8.4 mmol/L underwent a standard 75 g two-hour oral glucose tolerance test (OGTT). Fasting plasma glucose and 2h post-glucose, serum high-density lipoprotein-cholesterol (HDL-C), and serum triglycerides (TG), aspartate aminotransferase (AST) and alanine aminotransferase (ALT) were measured at baseline on a Roche Modular P Chemistry Analyzer (Roche Diagnostics). Low-density lipoprotein cholesterol (LDL-C) was estimated using the Friedewald’s formula when TG < 4.5 mmol/L. High sensitivity C-reactive protein (CRP) was measured using a Roche Modular P Chemistry Analyzer using the immunoturbidimetric method (Roche Diagnostics Corporation, IN). The study was approved by the institutional review boards at each field center, and all subjects provided written informed consent.

### HCHS/SOL clinical outcomes

eGFR was estimated from the serum creatinine-based Chronic Kidney Disease Epidemiology Collaboration equation ^18^. Prevalent CKD was defined based on an eGFR <60 ml/min/1.73 m^2^ at baseline and microalbuminuria was defined by a urine albumin to creatinine ratio [UACR] ≥ 30 mg/g at baseline. Other outcomes were diabetes, defined by a fasting glucose ≥126 mg/dl (7 mmol/L) or use of diabetes medications; prediabetes, defined by a fasting glucose of 100-125 mg/dL (5.6–7.0 mmol/L) and/or a HbA1c of 5.7–6.4% (39–46 mmol/mol); hypertension, defined by a blood pressure ≥130/80 mm Hg; and cardiovascular outcomes, including self-reported prevalent stroke, CAD and myocardial infarction. Incident CKD at visit 2 was defined by an eGFR <60 ml/min/1.73 m^2^ and an eGFR decline ≥1 ml/min per year, or UACR ≥30 mg/g at visit 2 among participants with an eGFR≥ 60 ml/min/1.73 m^2^ and a UACR <30 mg/g at baseline. For incident diabetes, we included only participants without diabetes at visit 1. No participant was taking SGLT2 inhibitors or incretin drugs.

### Genotyping

Participants were genotyped using the Illumina HumanOmni2.5-8v1-1 array as previously described^19^. Standard quality control was applied for variant- and sample-level quality filters. Principal components (PCs) were calculated using PC-AiR ^20^. The PCs showed genetic differentiation within and among six self-identified background groups (Cuban, Dominican, Puerto Rican, Mexican, and Central and South American). We removed 56 individuals with missing genetic background group. Imputation was performed using the TOPMed freeze 8 reference panel from the TOPMed imputation server ^21^, with Eagle v2.4 for phasing and Minimac4 for imputation ^22, 23^. We retained variants that had an imputation R^2^ ≥ 0.3.

### Identifying type 2 diabetes vs type 1 diabetes using a GRS

To exclude type 1 diabetes among HCHS/SOL participants, we calculated the type 1 diabetes genetic risk score 2 (GRS2) developed by Sharp et al., which includes 67 single nucleotide polymorphisms (SNPs)^11^. The GRS2 distinguishes type 1 diabetes from type 2 diabetes based on a bimodal distribution, using an approximate cutoff point of 13.5 that includes 87.8% of patients with type 2 diabetes and excludes 82.7% of type 1 diabetes patients^11^. Four SNPs (rs9405117, rs41295121, rs540653847, and rs1281943) were missing in the HCHS/SOL data. To test for unimodality, we ran Hartigan’s dip test. A p-value <0.05 indicates a non-unimodal distribution ^24^.

### Calculation of the FI clusters

We used the published weights for 110 SNPs to calculate seven FI cluster-specific PS^15^. Some SNPs contributed to multiple clusters. The alleles in these cluster-specific PS were aligned for increasing insulin effects. PS clusters were defined as Preserved IS, Elevated IS, Proinsulin, Adiposity-driven Hyperinsulinemia, Visceral Adiposity, Lipodystrophy and Liver/Lipid. For each FI cluster, the number of effect alleles for increasing FI at variants were multiplied by their respective multi-ancestry weights and summed across cluster variants to calculate each cluster-specific PS.

### Statistical analysis of FI PS clusters

We first tested each of the cluster-specific PS for associations with 19 traits used to cluster the FI SNPs: (a) among people without diabetes: fasting glucose, 2-hour glucose, HbA1c, HOMA-B and (b) overall sample: height, waist and hip circumference, waist/hip ratio, body mass index (BMI), HDL cholesterol, LDL cholesterol, triglycerides, body fat %, liver enzymes (AST, ALT), heart rate, PR interval, CRP, and urine albumin. We used linear mixed-effects models adjusted for age, sex, genetic group, center and five PCs, and random effects for census tract, household, and genetic relatedness to account for study design. Analyses for waist circumference, hip circumference, and waist/hip ratio were adjusted for BMI, while AST and ALT were natural log transformed. Associations with a p-value <3.8 x 10^-4^ were considered significant based on 133 tests (19 traits by seven FI clusters).

We then tested the association of each cluster with prediabetes, type 2 diabetes and insulin treatment use. The analysis for insulin use was only tested among individuals with type 2 diabetes, and individuals with diabetes were excluded from the prediabetes analyses. We also tested additional liver measures that were not used in the original clustering):Fibrosis-4 (FIB-4) index for liver fibrosis ^25^, hepatic steatosis index (HSI) ^26^, fatty liver index (FLI) ^25^ and non-alcoholic fatty liver disease liver fibrosis score (NAFLD−LFS) ^27^. FIB-4 and NAFLD-LFS were natural log transformed. We used the same linear mixed-effects models described above for clustering traits.

We then tested associations of each PS cluster with ten outcomes: CKD, CAD, stroke, myocardial infarction, hypertension, prediabetes, type 2 diabetes, insulin use, eGFR, and albuminuria. For the eGFR analysis, we used a linear mixed-effects model. For all categorical outcomes, we used logistic mixed-effects models. Models for CKD, eGFR, albuminuria, CAD, stroke, myocardial infarction, and hypertension were run separately by type 2 diabetes status, as well as jointly adjusting for type 2 diabetes status. The Bonferroni-adjusted significance level for all secondary outcomes was considered to be 5.1 x 10^-4^ (98 tests, 21 tests for three dysglycemic outcomes and seven FI clusters, and 49 tests for seven outcomes and seven FI clusters, and 28 for four liver traits by seven FI clusters).

### Analysis of measures of insulin homeostasis

#### MACAD and HTN-IR study description and genotyping

MACAD and HTN-IR cohorts are two similar cohorts of Mexican American families recruited from the Los Angeles area, as described previously ^28, 29^. We analyzed 1,316 individuals who did not have diabetes and who had undergone the euglycemic hyperinsulinemic clamp procedure, yielding direct measures of insulin sensitivity and insulin clearance. These participants were free of major medical illness, and none were taking glucocorticoids or other drugs that could affect glucose homeostasis. MACAD/HTN-IR participants were genotyped using the Illumina HumanOmniExpress Beadchip ^30^, with subsequent imputation using 1000 Genomes phase 3. Studies in MACAD/HTN-IR were approved by Human Subjects Protection Institutional Review Boards at UCLA, the University of Southern California, the Lundquist Institute, and Cedars-Sinai Medical Center. All subjects gave written informed consent prior to participation.

#### Insulin homeostasis traits

We analyzed insulin clearance, insulin secretion, insulin sensitivity, and disposition index. Insulin sensitivity and insulin clearance were measured in 1,316 non-diabetic individuals from the MACAD and HTN-IR cohorts via the euglycemic hyperinsulinemic clamp ^30^. The glucose infusion rate (M value) during the last 30 minutes of steady-state glucose and insulin levels during the clamp reflects glucose uptake by all tissues (mainly insulin-mediated glucose uptake in muscle) and is directly correlated with tissue insulin sensitivity. The metabolic clearance rate of insulin (MCRI) was calculated as the insulin infusion rate divided by the insulin concentration during the steady state of the euglycemic clamp ^31^. Multipoint OGTT was available in participants from MACAD (n=512). In those subjects, we calculated acute insulin secretion as the area under the curve (AUC) for insulin from baseline to 30 min over the corresponding AUC for glucose (AUC-Ins30/AUC-Glu30), which is highly correlated with first phase insulin secretion from the intravenous glucose tolerance test (r = 0.7) ^32^. This AUC-based insulin secretion measure has been found to have a hyperbolic relationship with insulin sensitivity, consistent with the relationship found when insulin secretion and insulin sensitivity are measured with gold standard physiologic tests ^33^. This relationship allows calculation of the disposition index, the product of insulin sensitivity and insulin secretion (M x AUC-Ins30/AUC-Glu30), which represents an index of insulin secretion that accounts for its degree of compensation for insulin resistance.

#### Statistical analysis

MACAD and HTN-IR participants were analyzed together as a single dataset. Phenotypes were log-transformed (disposition index) or square root-transformed (M value, MCRI, AUC-Ins30/AUC-Glu30) to normalize their distributions. We constructed cluster-specific PSs using SNPs and weights from Sevilla-Gonzalez et al.^15^. Two SNPs (rs84473 and rs8176749) were not available and rs84473 was replaced with a proxy, rs132941 (r^2^=0.92). Association between cluster-specific PS and phenotypes were evaluated using the general estimating equations method ^34^ assuming an exchangeable correlation structure and using the sandwich estimator of the variance to account for familial correlation. Cluster-specific PS association analyses were adjusted for age, sex, BMI, and four PCs to account for genetic variation in the data. Significance was considered a p-value < 0.002 based on testing seven clusters and four traits.

### Data and Resource Availability

The HCHS/SOL genotype and phenotype data is available at dbGap (phs000810.v1.p1). All codes used to define pPS are available in the supplemental material.

## Results

### Descriptive characteristics

HCHS/SOL participants mean age was 46 years, 59% were female, the average BMI was 29.8 kg/m^2^ and 20% had diabetes (**Table S1**). The prevalence of CKD was 3% (n=322), CAD was 5% (n=639), stroke was 1% (n=184), myocardial infarction was 2% (n=279), hypertension was 25% (n=3,158), and albuminuria was 10% (n=1,299). The distribution of clinical and biomarker traits used for clustering is shown in **Table S2**, and the distribution of liver biomarkers is shown in **Table S3**.

### Results for the type 1 diabetes GRS2

The type 1 diabetes GRS2 applied to the HCHS/SOL cohort had a unimodal distribution that closely matched the location and range of the type 2 diabetes individuals (**Figure S1**). The Hartigan’s dip test for unimodality p-value was 0.99, confirming no indication of a bimodal distribution. Therefore, all participants with diabetes were considered type 2 diabetes.

### Validation of FI clusters with clustering traits in HCHS/SOL Hispanics/Latinos

We first assessed whether FI clusters were associated with the cardiometabolic traits used to generate the clusters. For the Liver/lipid, Adiposity-driven Hyperinsulinemia, Lipodystrophy, and Preserved IS clusters, the results were concordant compared to associations in the published study that derived these clusters (**Figure 1; Tables S4-5**). The Adiposity-driven Hyperinsulinemia cluster was positively associated with increased BMI, waist to hip ratio, body fat %, natural log ALT, and HOMA-B. Conversely, the Lipodystrophy cluster was negatively associated with hip circumference, BMI, body fat % and HDL-cholesterol but positively associated with triglycerides and natural log ALT. Furthermore, the Liver/Lipid cluster was negatively associated with triglycerides, heart rate, CRP, and 2-hour glucose, and positively associated with fasting glucose. The Preserved IS cluster was negatively associated with 2-hour glucose.

**Figure 1.**
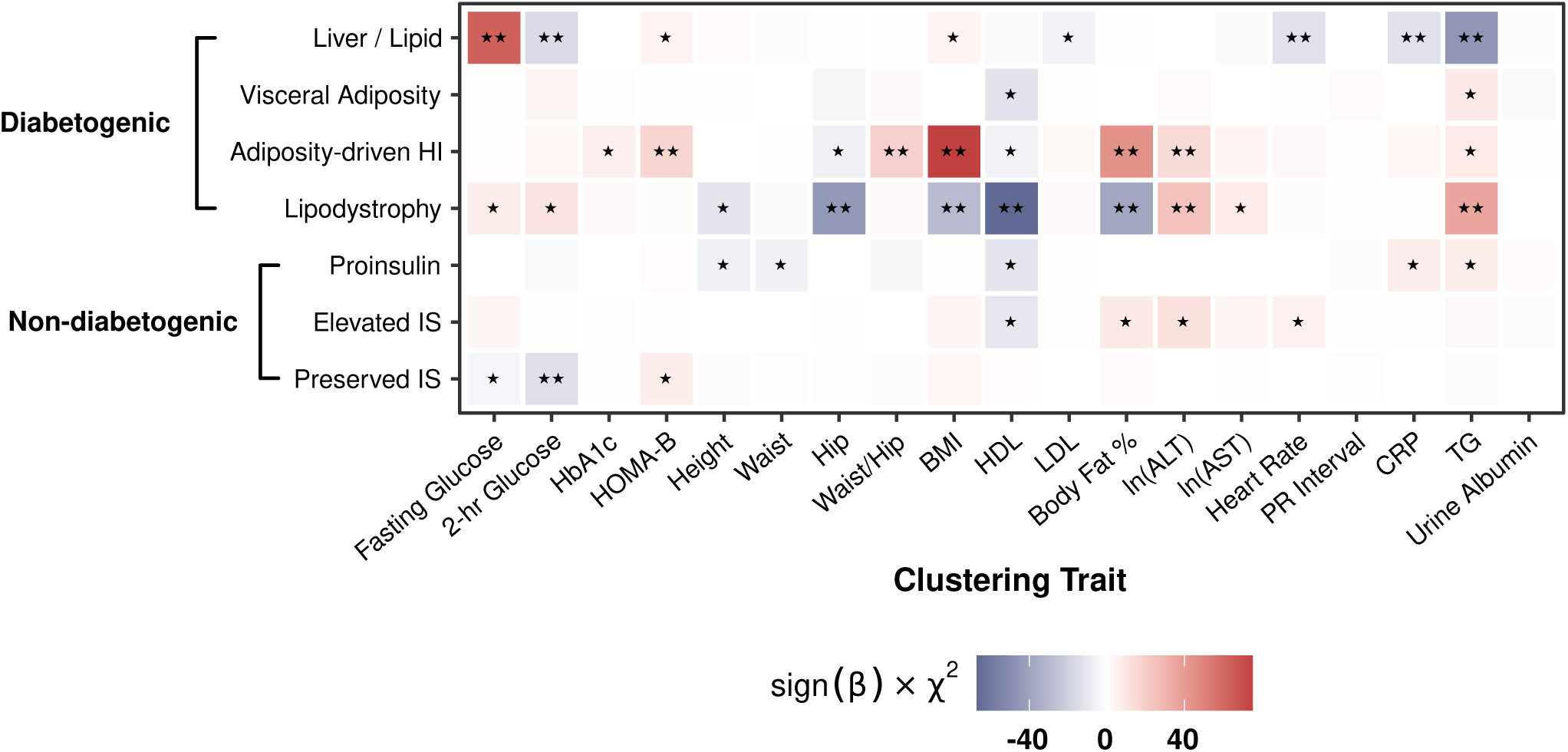
Heatmap showing strength of the associations between cluster PS and insulin clustering traits in the Hispanic Community Healthy Study / Study of Latinos. Only traits used in the clustering approach are shown. The Bonferroni-adjusted significance level was 3.8 × 10^−4^. One star means that the corresponding p-value is < 0.05, two stars means the result was significant at the Bonferroni-adjusted significance level. IS - insulin secretion; HI - hyperinsulinemia; FI - fasting insulin; OR - odds ratio; CI - confidence interval; HOMA-B - homeostasis model assessment of beta-cell function; BMI - body mass index; HDL - high- density lipoprotein; LDL - low-density lipoprotein; AST - aspartate aminotransferase; ALT - alanine aminotransferase; CRP – c-reactive protein; TG – triglycerides.

### Results for FI clusters with type 2 diabetes and prediabetes in HCHS/SOL

Consistent with the published findings, we reproduced a negative association of the Preserved IS (p-value = 4.0 x 10^-8^) with type 2 diabetes (**Figure 2; Table S6**) and the Lipodystrophy cluster-specific PS association with type 2 diabetes (p-value = 3.5 x 10^-5^), which were consistent with prior publication (**Table S7**). The FI cluster-specific PSs were not associated with insulin use among participants with type 2 diabetes. Associations of cluster-specific PS were not significant for prediabetes.

**Figure 2.**
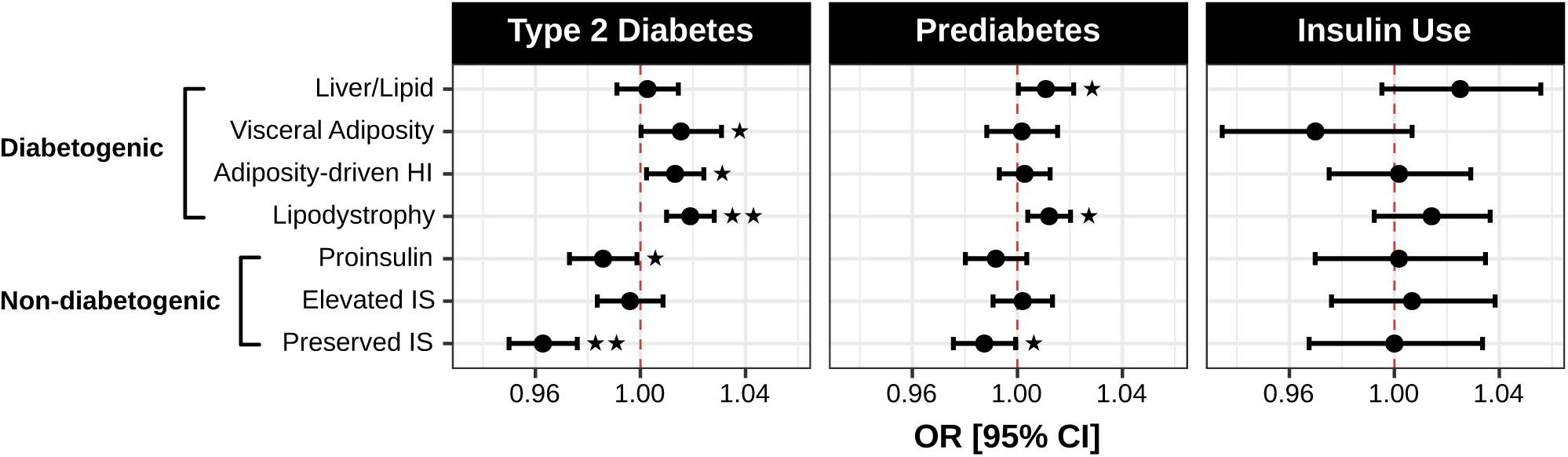
Forest plot of the associations between cluster PS and dysglycemia outcomes. The Bonferroni-adjusted significance level for these analyses was 5.1 x 10^-4^. One star means that the corresponding p-value is < 0.05, two stars means the result was significant at the Bonferroni-adjusted significance level.

### Results for FI clusters with liver fat and fibrosis

We also assessed associations with measurements of liver fat and fibrosis. The Elevated IS cluster was associated with the fatty liver index (p-value = 0.001) but not the hepatic steatosis index (**Table S8, Figure 3**). The Liver/Lipid cluster was associated with the natural log of NAFLD-LFS (p-value = 1.5 ×10^−4^)(**Table S9**). Additionally, the Adiposity-driven HI cluster was significantly positively associated with both hepatic steatosis index (p-value = 2.2 ×10^−17^) and fatty liver index (p-value = 1.7 ×10^−14^) (**Table S9**).

**Figure 3.**
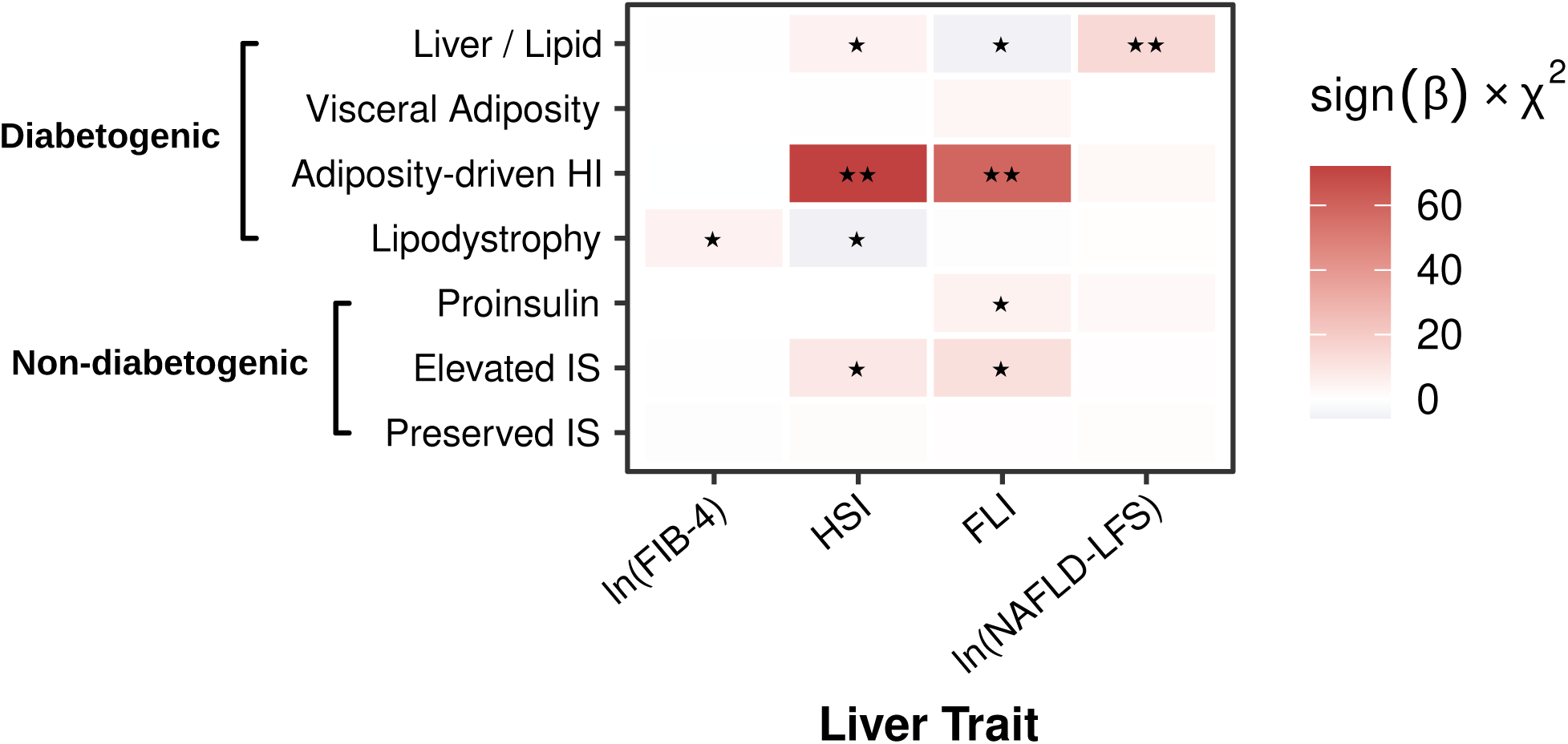
Heatmap showing strength of the associations between cluster PS and liver traits in the Hispanic Community Healthy Study / Study of Latinos. The Bonferroni-adjusted significance level was 5.1 x 10^-4^. One star means that the corresponding p-value is < 0.05, two stars means the result was significant at the Bonferroni-adjusted significance level. FI - fasting insulin; HI - hyperinsulinemia; FIB-4 - fibrosis-4 score; HSI - hepatic steatosis index; NAFLD-LFS - non-alcoholic fatty liver disease liver fat score. IS - insulin secretion; HI - hyperinsulinemia; FI - fasting insulin; OR - odds ratio; CI - confidence interval.

### Results for cardiovascular and kidney outcomes in HCHS/SOL

We then assessed the association of FI cluster-specific PS with cardiovascular and kidney outcomes. There were no significant associations with cardiovascular disease and hypertension (**Figure S2; Table S10**). For kidney outcomes, the Liver/Lipid cluster-specific PS was significantly associated with lower eGFR (p-value = 5.2 x 10^-5^)(**Figure 4; Table S11**). Associations with albuminuria and CKD were not significant for cluster-specific PS.

**Figure 4.**
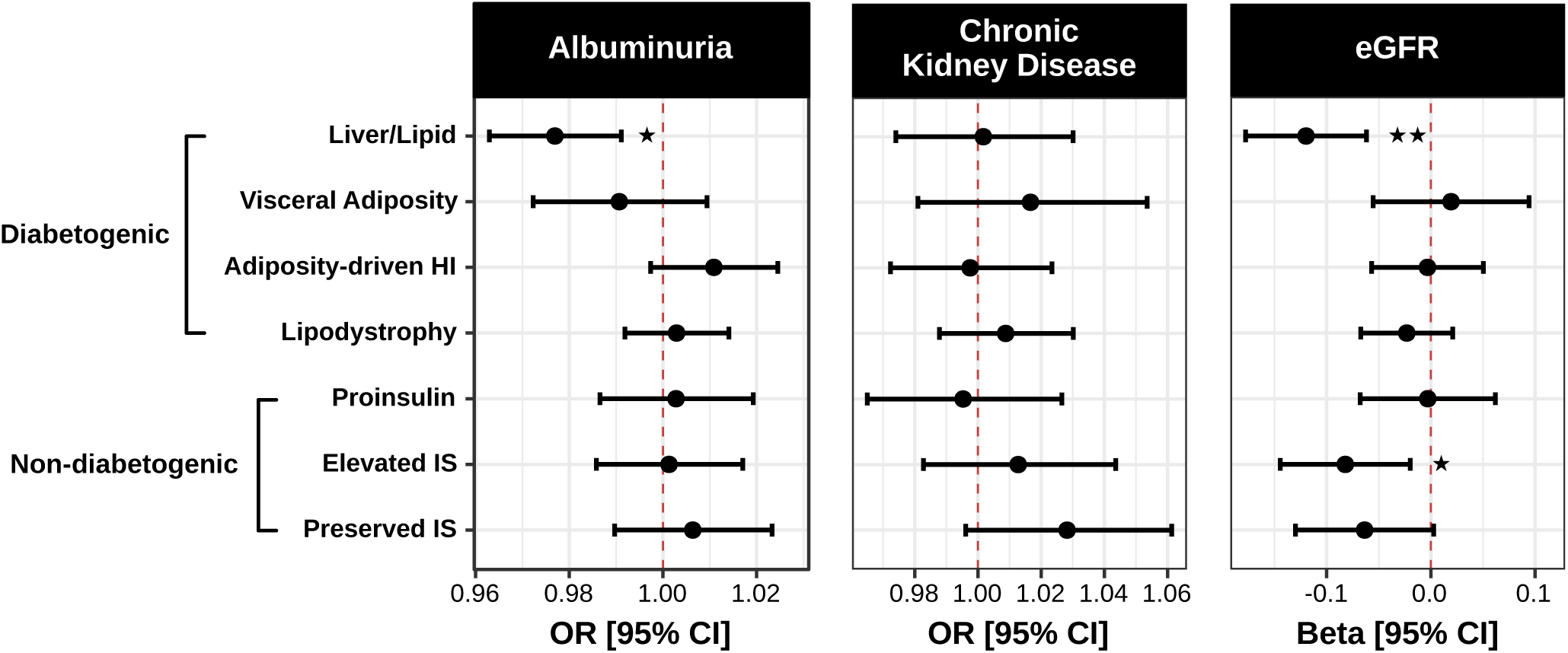
Forest plot of the associations between cluster PS and kidney outcomes. The Bonferroni-adjusted significance level for these analyses was 5.1 × 10^−4^. One star means that the corresponding p-value is < 0.05, two stars means the result was significant at the Bonferroni-adjusted significance level. CKD - chronic kidney disease; eGFR - estimated glomerular filtration rate; IS - insulin secretion; HI – hyperinsulinemia; OR - odds ratio; CI - confidence interval.

### Results for measures of insulin homeostasis in MACAD and HTN-IR

The participant characteristics and the distribution of insulin indexes are shown in **Table S12**. Association of the FI clusters with insulin homeostasis measures are displayed in **Figure S3** and **Tables S13** and **S14**. Several FI clusters were associated with insulin indexes but at a nominal level. The Elevated IS cluster-specific PS was significantly associated with log disposition index (p-value = 0.0015).

## Discussion

This study contributes new knowledge on FI genetic cluster associations with prediabetes, liver outcomes, and measures of insulin homeostasis using detailed physiologic phenotyping (euglycemic hyperinsulinemia clamp and OGTT) in Hispanics/Latinos, a population at high risk for type 2 diabetes and cardiometabolic complications. We first confirmed that HCHS/SOL Hispanics/Latinos with diabetes were more likely to have type 2 diabetes versus type 1 diabetes using a type 1 diabetes PS. We then tested FI cluster PSs and were able to reproduce patterns of association of these clusters related to distinct increased insulin mechanisms with cardiometabolic traits. Therefore, we validated for the first time FI cluster-cardiometabolic trait associations in Hispanics/Latinos, providing support for the relevance of these clusters in this population.

Our findings include positive associations of the Adiposity-driven Hyperinsulinemia and inverse associations of the Lipodystrophy cluster-specific PS with obesity measures of BMI and body fat %. The Lipodystrophy cluster-specific PS was associated with type 2 diabetes in our study. Sevilla-González et al. described an association of the Liver/Lipid cluster with type 2 diabetes, which we could not reproduce. Among the non-diabetogenic clusters, the Preserved IS cluster-specific PS was negatively associated with type 2 diabetes, in agreement with previously reported associations. There were no significant associations with prediabetes, or associations with insulin use among participants with diabetes.

Our main findings were significant associations of with liver measures, which have not been previously reported. The Adiposity-driven HI cluster-specific PS was associated with measures of liver fat, including the fatty liver index and the hepatic steatosis index. In contrast, the Liver/Lipid cluster-specific PS was associated with measures of liver fibrosis. Overall, these results provide insights into distinct liver outcomes for two diabetogenic hyperinsulinemia clusters related to liver fat accumulation and liver fibrosis. Because these associations are from biomarker-derived liver indexes, further studies are needed to assess the long-term clinical liver outcomes related to these FI clusters.

Our study also found a significant association of the Liver/Lipid cluster-specific PS with low eGFR. These results are concordant with findings from the original publication that showed associations of the Liver/Lipid cluster-specific PS with reduced lower eGFR and CKD ^15^. However, Sevilla-González et al. described additional associations of non-diabetogenic Preserved IS and Elevated IS clusters with CKD, which were not significant in our study. These kidney outcome findings will need to be further investigated in large longitudinal studies to better characterize CKD patterns and disease progression related to FI clusters, and experimental studies may uncover potential mechanisms and causal relationships, if associations are confirmed.

To gain insight into the physiologic mechanisms underlying the seven FI clusters, we assessed their association with measures of insulin sensitivity, insulin secretion, insulin clearance, and disposition index. We showed that the increased insulin secretion seen with the Elevated IS cluster could represent appropriate compensation or improved beta-cell function, a notion supported by increased disposition index. Disposition index, a measure of the beta-cell’s ability to compensate for insulin resistance, is a major determinant of diabetes risk, with higher levels of disposition index indicating lower risk of diabetes. For other clusters, associations were not significant.

Our study is limited to FI clusters previously developed, which used SNPs from GWAS of FI from individuals of European ancestry. Given Hispanics/Latinos have admixture of European, African and Native American ancestries, the contribution of SNPs from African and Amerindian ancestries were not included in the cluster development. Future studies including multi-population GWAS to develop FI clusters may improve prediction of clinical outcomes, especially cardiovascular outcomes in Hispanics/Latinos, which we were not able to identify. Additional limitations are the small number of cardiovascular events in our study, which were self-reported. A limitation to euglycemic hyperinsulinemic clamp measures of insulin sensitivity and insulin clearance is that they measure these traits on a whole-body level, such that organ-specific measures (e.g., hepatic insulin clearance vs kidney insulin clearance) are not available.

### Conclusion

This large study of Hispanics/Latinos validated cluster-specific PS clusters of FI and associations of the Lipodystrophy cluster-specific PS with type 2 diabetes. It also identified associations of the Liver/Lipid cluster-specific PS with liver fibrosis and low eGFR, and the Adiposity-driven HI with liver fat indexes. These findings support potential hyperinsulinemia mechanisms contributing to liver and kidney diseases. They also demonstrate that insulin has a quantifiable heterogeneous genetic foundation related to type 2 diabetes and other outcomes in Hispanics/Latinos that may identify mechanisms for disease risk to improve patient outcomes.

## Data Availability

The HCHS/SOL data is available through the dbGap access phs000810.v2.p2.

## List of abbreviations

AUC: area under the curve
ALT: alanine aminotransferase
AST: aspartate aminotransferase
BMI: body mass index
CAD: coronary artery disease
CKD: chronic kidney disease
CRP: high sensitivity C-reactive protein
FI: fasting insulin
HCHS/SOL: Hispanic Community Healthy Study/Study of Latinos
HDL-C: serum high-density lipoprotein-cholesterol
LDL-C: low-density lipoprotein cholesterol
MACAD: Mexican American Coronary Artery Disease
HTN-IR: Hypertension-Insulin Resistance
OGTT: oral glucose tolerance test
PC: principal component
SNP: single nucleotide polymorphism
TG: serum triglycerides
UACR: urine albumin to creatinine ratio
FIB-4: Fibrosis-4 index
HIS: hepatic steatosis index
FLI: fatty liver index
NAFLD-LFS: non-alcoholic fatty liver disease liver fibrosis score
MCRI: metabolic clearance rate of insulin.

## Acknowledgements

**Funding.** This study was also supported by the National Institutes of Health grants R01-DK117445, R01-HL163972, R01-HG013163, and K26-DK138425 to NF, and U01-DK140761 to JIR and MOG. MOG was supported by the Eris M. Field Chair in Diabetes Research. The Hispanic Community Health Study/Study of Latinos is a collaborative study supported by contracts from the National Heart, Lung, and Blood Institute (NHLBI) to the University of North Carolina (HHSN268201300001I / N01-HC-65233), University of Miami (HHSN268201300004I / N01-HC-65234), Albert Einstein College of Medicine (HHSN268201300002I / N01-HC-65235), University of Illinois at Chicago (HHSN268201300003I / N01-HC-65236 Northwestern Univ), and San Diego State University (HHSN268201300005I / N01-HC-65237). The following Institutes/Centers/Offices have contributed to the HCHS/SOL through a transfer of funds to the NHLBI: National Institute on Minority Health and Health Disparities, National Institute on Deafness and Other Communication Disorders, National Institute of Dental and Craniofacial Research, National Institute of Diabetes and Digestive and Kidney Diseases, National Institute of Neurological Disorders and Stroke, NIH Institution-Office of Dietary Supplements. APM acknowledges support from the National Institute for Health and Care Research (NIHR) Manchester Biomedical Research Centre (NIHR203308) and the British Heart Foundation (BHF) Research Excellence Award to the University of Manchester (RE/24/130017). The views expressed are those of the authors and not necessarily those of the NIHR or the Department of Health and Social Care. The study was supported in part by the National Center for Advancing Translational Sciences, CTSI grant UL1TR001881, the National Institute of Diabetes and Digestive and Kidney Disease Diabetes Research Center (DRC) grant DK063491 to the Southern California Diabetes Endocrinology Research Center, Hypertension and Insulin Resistance (HTN-IR) contracts R01-HL067974, R01-HL-55005, R01-HL 067974, U01DK140761, HL-088457 Mexican-American Coronary Artery Disease (MACAD), DK-079888 (work related to insulin clearance in HTN-IR and MACAD).

## Author’s contributions

N.F. and K.M.R. conceived the study and wrote the first draft of the manuscript. K.M.R., Ji.C., N.F., A.P.M. performed statistical analyses or supervised statistical methods. M.O.G. supervised analysis of the MACAD and HTN-IR studies. All authors edited the manuscript and approved the final version of the manuscript.

## Availability of data and materials

The HCHS/SOL data is available through the dbGap access phs000810.v2.p2.

## Supplemental Material

Tables S1-S14

Figures S1-S3

## References

1. Center for Disease Control. CDC National Diabetes Statistics Report. 2026.

2. American Diabetes Association Professional Practice Committee for D. 2. Diagnosis and Classification of Diabetes: Standards of Care in Diabetes-2026. Diabetes Care. 2026;49:S27–S49.

3. National Center for Health Statistics. National Health Interview Survey. Percentage of diagnosed diabetes for adults aged 18 and over, United States, 2019—2024. National Health Interview Survey.

4. Lee CJ, Bergman BK, Gou R, Williamson S and Boye KS. Prevalence, Demographic and Clinical Characteristics of Individuals with Early Onset Type 2 Diabetes in the USA: an NHANES Analysis 1999-2020. Diabetes Ther. 2025;16:2123–2136.

5. Glantz NM, Duncan I, Ahmed T, Fan L, Reed BL, Kalirai S and Kerr D. Racial and Ethnic Disparities in the Burden and Cost of Diabetes for US Medicare Beneficiaries. Health Equity. 2019;3:211–218.

6. Rowley WR, Bezold C, Arikan Y, Byrne E and Krohe S. Diabetes 2030: Insights from Yesterday, Today, and Future Trends. Popul Health Manag. 2017;20:6–12.

7. Kuusisto J, Mykkanen L, Pyorala K and Laakso M. Hyperinsulinemic microalbuminuria. A new risk indicator for coronary heart disease. Circulation. 1995;91:831–7.

8. Despres JP, Lamarche B, Mauriege P, Cantin B, Dagenais GR, Moorjani S and Lupien PJ. Hyperinsulinemia as an independent risk factor for ischemic heart disease. N Engl J Med. 1996;334:952–7.

9. Nilsson P, Nilsson JA, Hedblad B, Eriksson KF and Berglund G. Hyperinsulinaemia as long-term predictor of death and ischaemic heart disease in nondiabetic men: The Malmo Preventive Project. J Intern Med. 2003;253:136–45.

10. Yki-Jarvinen H. Fat in the liver and insulin resistance. Ann Med. 2005;37:347–56.

11. Sharp SA, Rich SS, Wood AR, Jones SE, Beaumont RN, Harrison JW, Schneider DA, Locke JM, Tyrrell J, Weedon MN, Hagopian WA and Oram RA. Development and Standardization of an Improved Type 1 Diabetes Genetic Risk Score for Use in Newborn Screening and Incident Diagnosis. Diabetes Care. 2019;42:200–207.

12. Suzuki K, Hatzikotoulas K, Southam L, Taylor HJ, Yin X, Lorenz KM, Mandla R, Huerta-Chagoya A, Melloni GEM, Kanoni S, Rayner NW, Bocher O, Arruda AL, Sonehara K, Namba S, Lee SSK, Preuss MH, Petty LE, Schroeder P, Vanderwerff B, Kals M, Bragg F, Lin K, Guo X, Zhang W, Yao J, Kim YJ, Graff M, Takeuchi F, Nano J, Lamri A, Nakatochi M, Moon S, Scott RA, Cook JP, Lee JJ, Pan I, Taliun D, Parra EJ, Chai JF, Bielak LF, Tabara Y, Hai Y, Thorleifsson G, Grarup N, Sofer T, Wuttke M, Sarnowski C, Gieger C, Nousome D, Trompet S, Kwak SH, Long J, Sun M, Tong L, Chen WM, Nongmaithem SS, Noordam R, Lim VJY, Tam CHT, Joo YY, Chen CH, Raffield LM, Prins BP, Nicolas A, Yanek LR, Chen G, Brody JA, Kabagambe E, An P, Xiang AH, Choi HS, Cade BE, Tan J, Broadaway KA, Williamson A, Kamali Z, Cui J, Thangam M, Adair LS, Adeyemo A, Aguilar-Salinas CA, Ahluwalia TS, Anand SS, Bertoni A, Bork-Jensen J, Brandslund I, Buchanan TA, Burant CF, Butterworth AS, Canouil M, Chan JCN, Chang LC, Chee ML, Chen J, Chen SH, Chen YT, Chen Z, Chuang LM, Cushman M, Danesh J, Das SK, de Silva HJ, Dedoussis G, Dimitrov L, Doumatey AP, Du S, Duan Q, Eckardt KU, Emery LS, Evans DS, Evans MK, Fischer K, Floyd JS, Ford I, Franco OH, Frayling TM, Freedman BI, Genter P, Gerstein HC, Giedraitis V, Gonzalez-Villalpando C, Gonzalez-Villalpando ME, Gordon-Larsen P, Gross M, Guare LA, Hackinger S, Hakaste L, Han S, Hattersley AT, Herder C, Horikoshi M, Howard AG, Hsueh W, Huang M, Huang W, Hung YJ, Hwang MY, Hwu CM, Ichihara S, Ikram MA, Ingelsson M, Islam MT, Isono M, Jang HM, Jasmine F, Jiang G, Jonas JB, Jorgensen T, Kamanu FK, Kandeel FR, Kasturiratne A, Katsuya T, Kaur V, Kawaguchi T, Keaton JM, Kho AN, Khor CC, Kibriya MG, Kim DH, Kronenberg F, Kuusisto J, Lall K, Lange LA, Lee KM, Lee MS, Lee NR, Leong A, Li L, Li Y, Li-Gao R, Ligthart S, Lindgren CM, Linneberg A, Liu CT, Liu J, Locke AE, Louie T, Luan J, Luk AO, Luo X, Lv J, Lynch JA, Lyssenko V, Maeda S, Mamakou V, Mansuri SR, Matsuda K, Meitinger T, Melander O, Metspalu A, Mo H, Morris AD, Moura FA, Nadler JL, Nalls MA, Nayak U, Ntalla I, Okada Y, Orozco L, Patel SR, Patil S, Pei P, Pereira MA, Peters A, Pirie FJ, Polikowsky HG, Porneala B, Prasad G, Rasmussen-Torvik LJ, Reiner AP, Roden M, Rohde R, Roll K, Sabanayagam C, Sandow K, Sankareswaran A, Sattar N, Schonherr S, Shahriar M, Shen B, Shi J, Shin DM, Shojima N, Smith JA, So WY, Stancakova A, Steinthorsdottir V, Stilp AM, Strauch K, Taylor KD, Thorand B, Thorsteinsdottir U, Tomlinson B, Tran TC, Tsai FJ, Tuomilehto J, Tusie-Luna T, Udler MS, Valladares-Salgado A, van Dam RM, van Klinken JB, Varma R, Wacher-Rodarte N, Wheeler E, Wickremasinghe AR, van Dijk KW, Witte DR, Yajnik CS, Yamamoto K, Yamamoto K, Yoon K, Yu C, Yuan JM, Yusuf S, Zawistowski M, Zhang L, Zheng W, Program VAMV, Raffel LJ, Igase M, Ipp E, Redline S, Cho YS, Lind L, Province MA, Fornage M, Hanis CL, Ingelsson E, Zonderman AB, Psaty BM, Wang YX, Rotimi CN, Becker DM, Matsuda F, Liu Y, Yokota M, Kardia SLR, Peyser PA, Pankow JS, Engert JC, Bonnefond A, Froguel P, Wilson JG, Sheu WHH, Wu JY, Hayes MG, Ma RCW, Wong TY, Mook-Kanamori DO, Tuomi T, Chandak GR, Collins FS, Bharadwaj D, Pare G, Sale MM, Ahsan H, Motala AA, Shu XO, Park KS, Jukema JW, Cruz M, Chen YI, Rich SS, McKean-Cowdin R, Grallert H, Cheng CY, Ghanbari M, Tai ES, Dupuis J, Kato N, Laakso M, Kottgen A, Koh WP, Bowden DW, Palmer CNA, Kooner JS, Kooperberg C, Liu S, North KE, Saleheen D, Hansen T, Pedersen O, Wareham NJ, Lee J, Kim BJ, Millwood IY, Walters RG, Stefansson K, Ahlqvist E, Goodarzi MO, Mohlke KL, Langenberg C, Haiman CA, Loos RJF, Florez JC, Rader DJ, Ritchie MD, Zollner S, Magi R, Marston NA, Ruff CT, van Heel DA, Finer S, Denny JC, Yamauchi T, Kadowaki T, Chambers JC, Ng MCY, Sim X, Below JE, Tsao PS, Chang KM, McCarthy MI, Meigs JB, Mahajan A, Spracklen CN, Mercader JM, Boehnke M, Rotter JI, Vujkovic M, Voight BF, Morris AP and Zeggini E. Genetic drivers of heterogeneity in type 2 diabetes pathophysiology. Nature. 2024;627:347–357.

13. Udler MS, Kim J, Marcin von G, Cole JB, Chiou J, on behalf of M, the I, Boehnke M, Laakso M, Atzmon G, Glaser B, Mercader JM, Gaulton K, Flannick J, Getz G and Florez JC. Type 2 diabetes genetic loci informed by multi-trait associations point to disease mechanisms and subtypes: A soft clustering analysis. PLoS Medicine. 2018;15.

14. Smith K, Deutsch AJ, McGrail C, Kim H, Hsu S, Mandla R, Schroeder PH, Westerman KE, Szczerbinski L, Majarian TD, Kaur V, Williamson A, Claussnitzer M, Florez JC, Manning AK, Mercader JM, Gaulton KJ and Udler MS. Multi-ancestry Polygenic Mechanisms of Type 2 Diabetes Elucidate Disease Processes and Clinical Heterogeneity. medRxiv. 2023.

15. Sevilla-Gonzalez M, Smith K, Wang N, Jensen AE, Litkowski EM, Kim H, DiCorpo DA, Hsu S, Cui J, Liu CT, Yu C, McNeil JJ, Lacaze P, Westerman KE, Chang KM, Tsao PS, Phillips LS, Goodarzi MO, Sladek R, Rotter JI, Dupuis J, Florez JC, Merino J, Meigs JB, Zhou JJ, Raghavan S, Udler MS and Manning AK. Heterogeneous effects of genetic variants and traits associated with fasting insulin on cardiometabolic outcomes. Nat Commun. 2025;16:2569.

16. Sorlie PD, Aviles-Santa LM, Wassertheil-Smoller S, Kaplan RC, Daviglus ML, Giachello AL, Schneiderman N, Raij L, Talavera G, Allison M, Lavange L, Chambless LE and Heiss G. Design and implementation of the Hispanic Community Health Study/Study of Latinos. Ann Epidemiol. 2010;20:629–41.

17. Thyagarajan B, Howard AG, Durazo-Arvizu R, Eckfeldt JH, Gellman MD, Kim RS, Liu K, Mendez AJ, Penedo FJ, Talavera GA, Youngblood ME, Zhao L and Sotres-Alvarez D. Analytical and biological variability in biomarker measurement in the Hispanic Community Health Study/Study of Latinos. Clin Chim Acta. 2016;463:129–137.

18. Inker LA, Schmid CH, Tighiouart H, Eckfeldt JH, Feldman HI, Greene T, Kusek JW, Manzi J, Van Lente F, Zhang YL, Coresh J, Levey AS and Investigators C-E. Estimating glomerular filtration rate from serum creatinine and cystatin C. N Engl J Med. 2012;367:20–9.

19. Conomos MP, Laurie CA, Stilp AM, Gogarten SM, McHugh CP, Nelson SC, Sofer T, Fernandez-Rhodes L, Justice AE, Graff M, Young KL, Seyerle AA, Avery CL, Taylor KD, Rotter JI, Talavera GA, Daviglus ML, Wassertheil-Smoller S, Schneiderman N, Heiss G, Kaplan RC, Franceschini N, Reiner AP, Shaffer JR, Barr RG, Kerr KF, Browning SR, Browning BL, Weir BS, Aviles-Santa ML, Papanicolaou GJ, Lumley T, Szpiro AA, North KE, Rice K, Thornton TA and Laurie CC. Genetic Diversity and Association Studies in US Hispanic/Latino Populations: Applications in the Hispanic Community Health Study/Study of Latinos. Am J Hum Genet. 2016;98:165–84.

20. Conomos MP, Miller MB and Thornton TA. Robust inference of population structure for ancestry prediction and correction of stratification in the presence of relatedness. Genet Epidemiol. 2015;39:276–93.

21. Sun Q, Liu W, Rosen JD, Huang L, Pace RG, Dang H, Gallins PJ, Blue EE, Ling H, Corvol H, Strug LJ, Bamshad MJ, Gibson RL, Pugh EW, Blackman SM, Cutting GR, O’Neal WK, Zhou YH, Wright FA, Knowles MR, Wen J, Li Y and Cystic Fibrosis Genome P. Leveraging TOPMed imputation server and constructing a cohort-specific imputation reference panel to enhance genotype imputation among cystic fibrosis patients. HGG Adv. 2022;3:100090.

22. Loh PR, Danecek P, Palamara PF, Fuchsberger C, Y AR, H KF, Schoenherr S, Forer L, McCarthy S, Abecasis GR, Durbin R and A LP. Reference-based phasing using the Haplotype Reference Consortium panel. Nat Genet. 2016;48:1443–1448.

23. Fuchsberger C, Abecasis GR and Hinds DA. minimac2: faster genotype imputation. Bioinformatics. 2015;31:782–4.

24. Hartigan JA and Hartigan PA. The Dip Test of Unimodality. The Annals of Statistics. 1985;13:70–84.

25. Sterling RK, Lissen E, Clumeck N, Sola R, Correa MC, Montaner J, M SS, Torriani FJ, Dieterich DT, Thomas DL, Messinger D, Nelson M and Investigators AC. Development of a simple noninvasive index to predict significant fibrosis in patients with HIV/HCV coinfection. Hepatology. 2006;43:1317–25.

26. Lee JH, Kim D, Kim HJ, Lee CH, Yang JI, Kim W, Kim YJ, Yoon JH, Cho SH, Sung MW and Lee HS. Hepatic steatosis index: a simple screening tool reflecting nonalcoholic fatty liver disease. Dig Liver Dis. 2010;42:503–8.

27. Kotronen A, Peltonen M, Hakkarainen A, Sevastianova K, Bergholm R, Johansson LM, Lundbom N, Rissanen A, Ridderstrale M, Groop L, Orho-Melander M and Yki-Jarvinen H. Prediction of non-alcoholic fatty liver disease and liver fat using metabolic and genetic factors. Gastroenterology. 2009;137:865–72.

28. Goodarzi MO, Guo X, Taylor KD, Quinones MJ, Samayoa C, Yang H, Saad MF, Palotie A, Krauss RM, Hsueh WA and Rotter JI. Determination and use of haplotypes: ethnic comparison and association of the lipoprotein lipase gene and coronary artery disease in Mexican-Americans. Genet Med. 2003;5:322–7.

29. Xiang AH, Azen SP, Raffel LJ, Tan S, Cheng LS, Diaz J, Toscano E, Henderson PC, Hodis HN, Hsueh WA, Rotter JI and Buchanan TA. Evidence for joint genetic control of insulin sensitivity and systolic blood pressure in hispanic families with a hypertensive proband. Circulation. 2001;103:78–83.

30. Palmer ND, Goodarzi MO, Langefeld CD, Wang N, Guo X, Taylor KD, Fingerlin TE, Norris JM, Buchanan TA, Xiang AH, Haritunians T, Ziegler JT, Williams AH, Stefanovski D, Cui J, Mackay AW, Henkin LF, Bergman RN, Gao X, Gauderman J, Varma R, Hanis CL, Cox NJ, Highland HM, Below JE, Williams AL, Burtt NP, Aguilar-Salinas CA, Huerta-Chagoya A, Gonzalez-Villalpando C, Orozco L, Haiman CA, Tsai MY, Johnson WC, Yao J, Rasmussen-Torvik L, Pankow J, Snively B, Jackson RD, Liu S, Nadler JL, Kandeel F, Chen YD, Bowden DW, Rich SS, Raffel LJ, Rotter JI, Watanabe RM and Wagenknecht LE. Genetic variants associated with quantitative glucose homeostasis traits translate to type 2 diabetes in Mexican Americans: the GUARDIAN (Genetics Underlying Diabetes in Hispanics) Consortium. Diabetes. 2015;64:1853–66.

31. DeFronzo RA, Tobin JD and Andres R. Glucose clamp technique: a method for quantifying insulin secretion and resistance. The American journal of physiology. 1979;237:E214–23.

32. Stancakova A, Javorsky M, Kuulasmaa T, Haffner SM, Kuusisto J and Laakso M. Changes in insulin sensitivity and insulin release in relation to glycemia and glucose tolerance in 6,414 Finnish men. Diabetes. 2009;58:1212–21.

33. Santos JL, Yevenes I, Cataldo LR, Morales M, Galgani J, Arancibia C, Vega J, Olmos P, Flores M, Valderas JP and Pollak F. Development and assessment of the disposition index based on the oral glucose tolerance test in subjects with different glycaemic status. Journal of physiology and biochemistry. 2016;72:121–31.

34. Zeger SL and Liang KY. Longitudinal data analysis for discrete and continuous outcomes. Biometrics. 1986;42:121–30.

